# Acute kidney injury in patients hospitalized with COVID-19 in Wuhan, China: A single-center retrospective observational study

**DOI:** 10.1101/2020.04.06.20055194

**Authors:** Guanhua Xiao, Hongbin Hu, Feng Wu, Tong Sha, Qiaobing Huang, Haijun Li, Jiafa Han, Wenhong Song, Zhongqing Chen, Zhenhua Zeng

## Abstract

**Background:** The kidney may be affected in coronavirus-2019 disease (COVID-19). This study assessed the predictors and outcomes of acute kidney injury (AKI) among individuals with COVID-19.

**Methods:** This observational study, included data on all patients with clinically confirmed COVID-19 admitted to Hankou Hospital, Wuhan, China from January 5 to March 8, 2020. Data were extracted from clinical and laboratory records. Follow-up was censored on March 8, 2020.

This is a single-center, retrospective, observational study. Patients clinically confirmed COVID-19 and admitted to Hankou Hospital, Wuhan, China from January 5 to March 8, 2020 were enrolled. We evaluated the association between changes in the incidence of AKI and COVID-19 disease and clinical outcomes by using logistic regression models.

**Results:** A total of 287 patients, 55 with AKI and 232 without AKI, were included in the analysis. Compared to patients without AKI, AKI patients were older, predominantly male, and were more likely to present with hypoxia and have pre-existing hypertension and cerebrovascular disease. Moreover, AKI patients had higher levels of white blood cells, D-dimer, aspartate aminotransferase, total bilirubin, creatine kinase, lactate dehydrogenase, procalcitonin, C-reactive protein, a higher prevalence of hyperkalemia, lower lymphocyte counts, and higher chest computed tomographic scores. The incidence of stage 1 AKI was 14.3%, and the incidence of stage 2 or 3 AKI was 4.9%. Patients with AKI had substantially higher mortality.

**Conclusions:** AKI is an important complication of COVID-19. Older age, male, multiple pre-existing comorbidities, lymphopenia, increased infection indicators, elevated D-dimer, and impaired heart and liver functions were the risk factors of AKI. AKI patients who progressed to stages 2 or 3 AKI had a higher mortality rate. Prevention of AKI and monitoring of kidney function is very important for COVID-19 patients.

**Trial registration:** NCT04316299(03/19/2020)

## Background

In December, 2019, a cluster of severe pneumonia cases of unknown cause emerged in Wuhan, China, with clinical presentations greatly resembling viral pneumonia[1]. Deep sequencing analysis from lower respiratory tract samples indicated a novel coronavirus, which was named 2019 novel coronavirus (2019-nCoV, then named COVID-19). Up to February 24, 2020, the total number of patients had risen sharply to 77,269 confirmed cases and 2,596 deaths cases (http://2019ncov.chinacdc.cn/2019-nCoV/). The number of critically ill patients is as high as 9915. The clinical spectrum of COVID-19 pneumonia ranges from non-severe to severe cases[2]. Previous studies have described the general epidemiological findings, clinical presentation, and clinical outcomes of patients of COVID-19 pneumonia[3–8]. Of note, latest data suggests that the kidney is the most vulnerable organ in COVID-19 patients besides the lungs, with evidence of acute kidney injury (AKI) in up to 37.5% of fatal cases of COVID-19 [3]. However, there is currently no published information characterizing AKI risk factors and outcomes in patients with COVID-19. In this study, we investigated AKI in patients admitted to Hankou Hospital in Wuhan with confirmed COVID-19 pneumonia and determined the factors associated with AKI in patients hospitalized with COVID-19 in order to identify factors that can be used for early identification of individuals who are at risk of developing AKI, so that they can be provided with intensive care treatment, thereby reducing the mortality and morbidity associated with AKI in patients with COVID-19.

## Methods

### Study design and participants

This single-center, retrospective, observational study was conducted at Hankou Hospital in Wuhan, China, which is a hospital designated to treat COVID-19 pneumonia patients. The criteria for suspected and confirmed cases are based on criteria previously established by WHO [7, 8] and patients who met the WHO criteria were included in our study. Patients who were younger than 18 years, who had undergone renal replacement therapy (RRT) before admission, or whose entire stay lasted for less than 48 hours were excluded from the analysis. The primary variable of interest was the incidence of AKI, and secondary outcome variables included the 28-day mortality (calculated from the day of admission), and the length of hospital stay. This study was approved by the National Health Commission of China and the Ethics Commission of Hankou Hospital (hkyy2020-005). The Ethics Commission of Hankou Hospital waived the requirement for obtaining informed consent.

### Data collection and definitions

We obtained epidemiological, demographic, clinical, laboratory, management, and outcome data from patients’ medical records using standardized data collection forms. Clinical outcomes were followed up to March 8, 2020. To analyze the relationship between AKI staging and clinical outcomes, a minimum follow-up time of 14 days was required, including follow-up of patients who were discharged before March 8, 2020. If data were missing from the records or clarification was needed, the researchers also directly communicated with patients or their families to ascertain epidemiological and symptom data. All data were checked by two physicians (GX and FZ). Initial investigations included a complete blood count, coagulation profile, and serum biochemical test (including renal and liver function, creatine kinase, lactate dehydrogenase (LDH), electrolytes, and 10 common respiratory pathogens. A description of the treatment procedures and measurements has been published previously[7].

All patients had a chest computed tomography (CT) scan on admission. All CT images were reviewed by two fellowship-trained cardiothoracic radiologists (GX and FW), with approximately 5 years of experience each, using the Electronic Medical Record System of Hankou Hospital. Images were reviewed independently, and final decisions were reached by consensus. In cases of disagreement between the two primary radiologist interpretations, a third fellowship-trained cardiothoracic radiologist with 10 years of experience (HL.) adjudicated the final decision. No negative control cases were examined, and no blinding occurred. The radiologists evaluated the initial CT scans and assigned quantitative scores for the characteristics using a recently published scoring system[9]. The total CT score was the sum of lung involvement (5 lobes, score 1-5 for each lobe, range, 0 none, 25 maximum) was determined[10].

Acute kidney injury (AKI) was identified and classified on the basis of the highest serum creatinine (Cr) level according to the Kidney Disease Improving Global Outcomes (KDIGO) definition and staging system[11].However, urine output is not routinely monitored in our hospital, which rendered this approach infeasible. Thus, we used increased creatinine alone for the diagnosis and staging of AKI. Besides, a missing baseline serum creatinine measurement is existed in some patients before admission. Two alternative methods were used to estimate the baseline creatinine value in order to make the diagnosis of AKI. The average serum creatinine method used non-AKI patient data to calculate the average serum creatinine value based on age and sex [12] and the estimated glomerular filtration rate (eGFR) method used an Modification of Diet in Renal Disease (MDRD) equation to determine the eGFR value based on age and sex (75 mL/min/1.73 m^2^)[11, 13]. Briefly, the AKI diagnostic criteria are: an increase in serum creatinine by ≥26.5 μmol/L within 48 hours, or an increase in serum creatinine to ≥1.5 times baseline in <7 days, using a known or presumed baseline value. The AKI Staging criteria are: **Stage 1**: Increase in serum creatinine to 1.5 to 1.9 times baseline, or increase in serum creatinine by ≥26.5 μmol/L. **Stage 2**: Increase in serum creatinine to 2.0 to 2.9 times baseline.

**Stage 3**: Increase in serum creatinine to 3.0 times baseline, or increase in serum creatinine to ≥353.6 μmol/L, or the initiation of renal replacement therapy.

### Statistical analysis

Statistical analysis was performed using SPSS software for Windows, version 20.0 (IBM Corp, Armonk, NY, USA). Descriptive analyses were performed for demographic, clinical, and laboratory data. Bivariate analyses were used to assess the association of AKI status and different parameters of interest. Continuous data, such as white blood cell count was converted into categorical variables (normal, low, or high levels). Multivariate analysis was not performed due to the small sample size. The odds ratio (OR), obtained by logistic regression, was used to measure the strength of the association of each factor with AKI. A P value <*0*.*05* was considered to indicate statistical significance. The date of onset of symptoms was used as the starting date. We conducted a death certificate search of medical records to determine whether any patients who were alive on discharge died subsequently, and included these data in the analysis when applicable.

## Results

During the study period, a total of 287 patients were admitted and met the case definition of clinically confirmed COVID-19 infection and the other inclusion criteria. The characteristics of the patients on admission overall, and according to AKI status are shown in **Table 1**. There were 55 patients with AKI and 232 patients without AKI. The incidence of AKI was 19.2% according to the average serum creatinine method. Compared to patients without AKI, patients with AKI were more likely to be male, were significantly older; and were more likely to have underlying comorbidities, including chronic renal insufficiency, hypertension, and cerebrovascular disease. Patients with AKI tended to have more severe pneumonia, and were more likely to have hypoxia and tachypnea. **Table 2** shows the results of laboratory tests performed on admission according to AKI status. Compared to patients without AKI, patients with AKI had higher white blood cell and neutrophil counts; higher levels of D-dimer, aspartate aminotransferase, total bilirubin, creatine kinase, LDH, procalcitonin (PCT), and C-reactive protein (CRP); and a higher prevalence of hyperkalemia. Compared to patients without AKI, patients with AKI had lower lymphocyte counts and lower serum albumin.

**Table 1.**
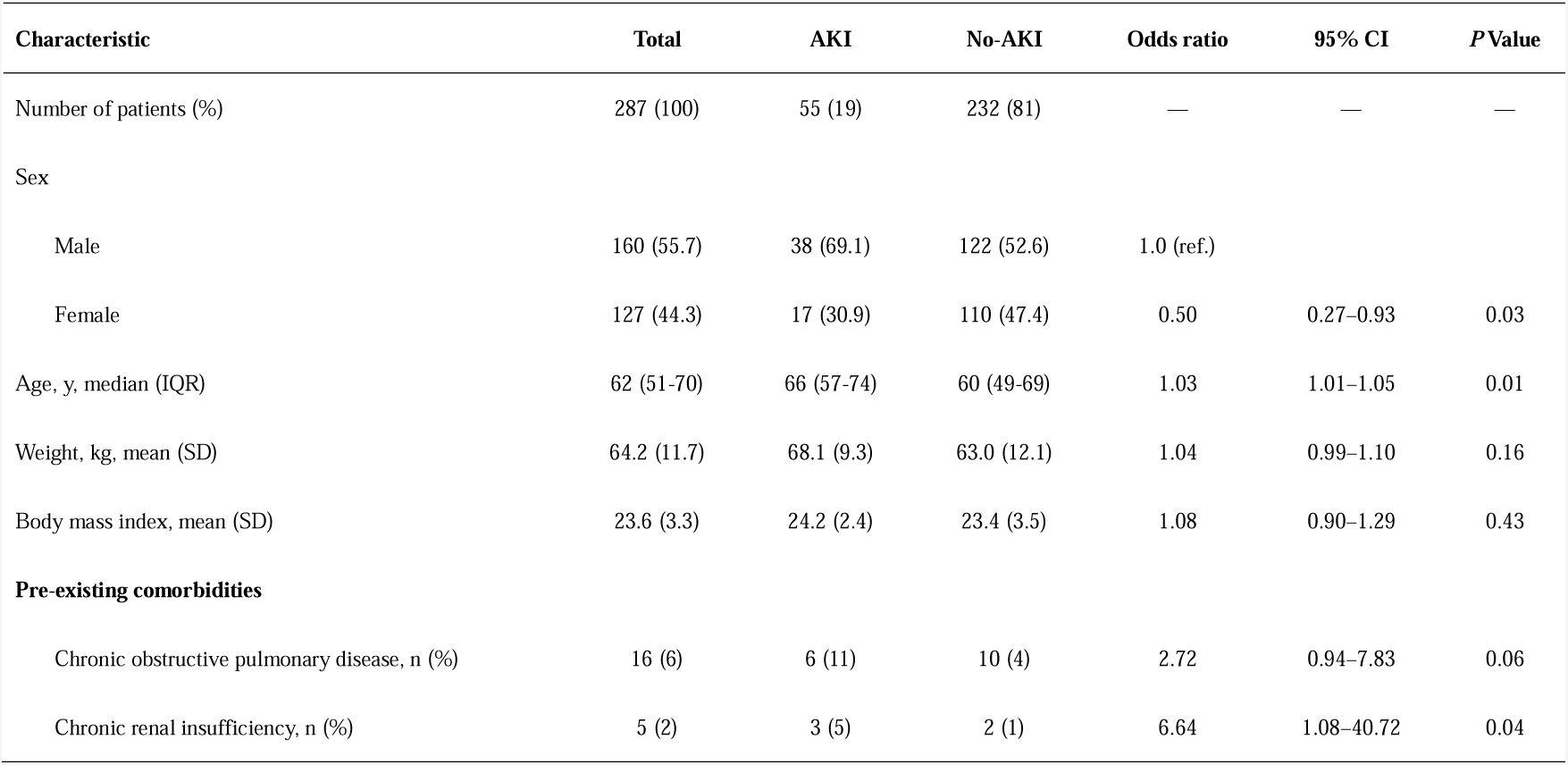
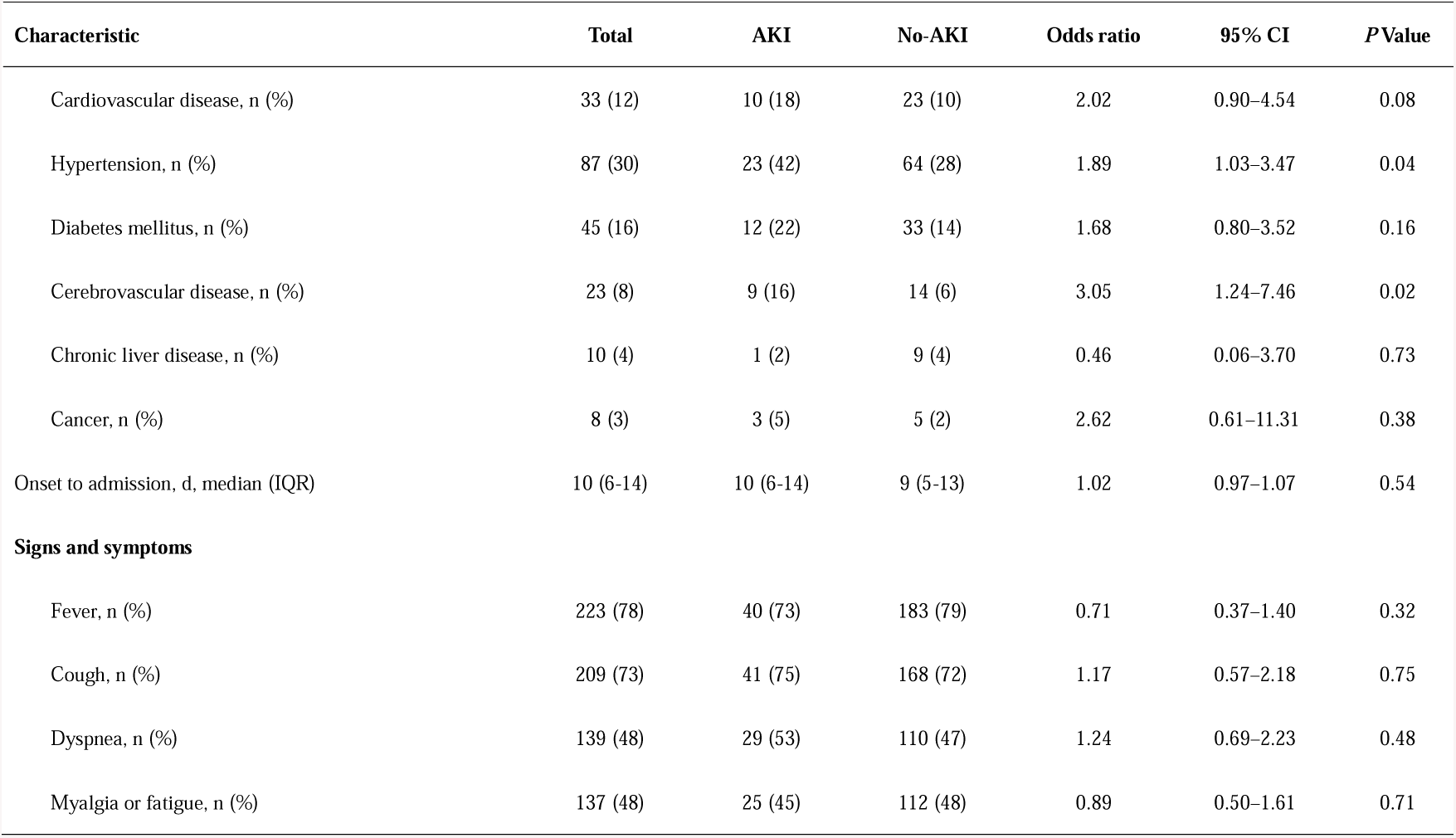
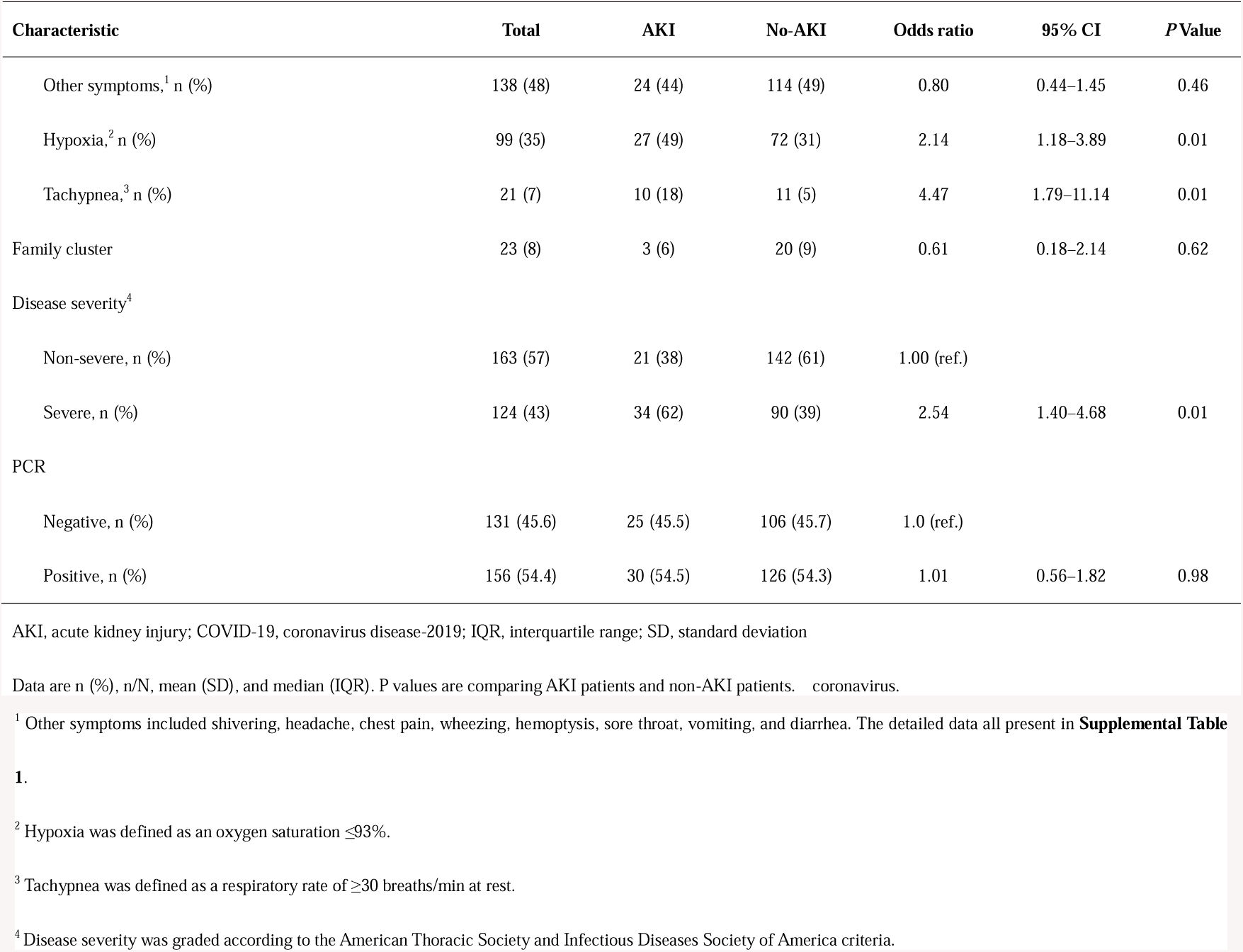
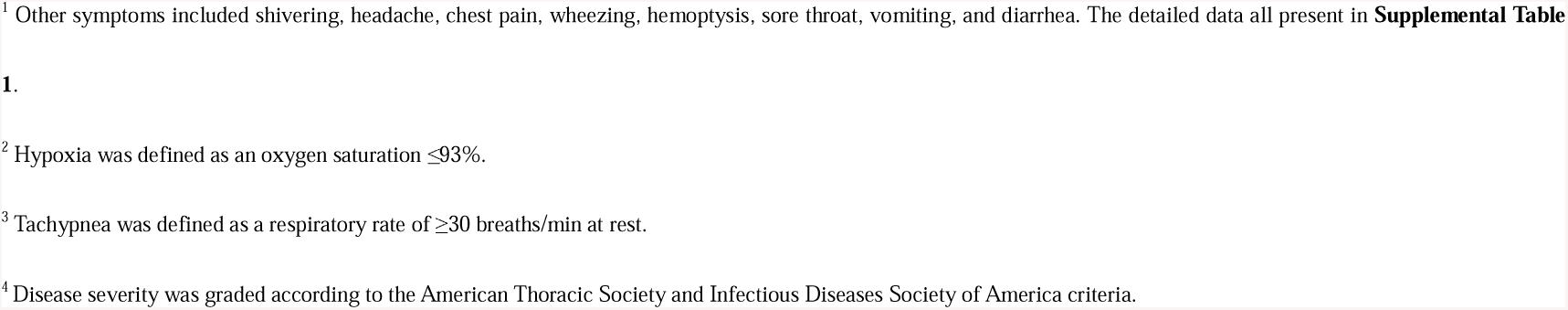
Characteristics of COVID-19 patients on admission to hospital according to acute kidney injury status

**Table 2.** Laboratory results of COVID-19 patients on admission to hospital according to acute kidney injury status

|  | Normal range | Total<br>(n = 287) | AKI<br>(n = 55) | Non-AKI<br>(n = 232) | *P* Value |
| --- | --- | --- | --- | --- | --- |
| White blood cell count ($\times 10^9/L$), median (IQR) | 4.0–10.0 | 5.0 (3.8–7.2) | 5.8 (4.3–9.5) | 4.9 (3.6–6.7) | <0.01 |
| <4, n (%) |  | 129 (45) | 9 (16) | 120 (52) |  |
| 4–10, n (%) |  | 131 (46) | 34 (62) | 97 (42) | <0.01 |
| >10, n (%) |  | 27 (9) | 12 (22) | 15 (6) |  |
| Neutrophil count ($\times 10^9/L$), median (IQR) | 1.8–6.3 | 3.6 (2.3–5.5) | 4.5 (3.0–8.5) | 3.4 (2.2–5.1) | <0.01 |
| Lymphocyte count ($\times 10^9/L$), median (IQR) | 1.1–3.2 | 0.8 (0.6–1.2) | 0.7 (0.4–1.0) | 0.9 (0.6–1.2) | <0.01 |
| Hemoglobin (g/L), mean (SD) | 120–150 | 128.3 (17.1) | 125 (20.0) | 129.1 (16.3) | 0.11 |
| Prothrombin time (s), mean (SD) | 11.0–16.0 | 15.3 (5.8) | 17.1 (11.7) | 14.9 (2.8) | 0.11 |
| Activated partial thromboplastin time (s), mean (SD) | 27.0–45.0 | 35.8 (7.3) | 37.7 (10.5) | 35.4 (6.3) | 0.05 |
| D-dimer (mg/mL), median (IQR) | 0.0–0.5 | 0.6 (0.2–2.0) | 0.9 (0.2–6.6) | 0.5 (0.2–1.6) | <0.01 |
| Alanine aminotransferase (U/L), median (IQR) | 0–40 | 25 (17–38) | 27 (18–43) | 24 (16–37) | 0.18 |
| Aspartate aminotransferase (U/L), median (IQR) | 0–40 | 27 (20–41) | 35 (23–50) | 26 (19–40) | <0.01 |
| Total bilirubin (μmol/L), median (IQR) | 3.4–20.5 | 8.9 (6.6–12.0) | 11.0 (8.1–16.0) | 8.6 (6.3–11.38) | <0.01 |
| Albumin (g/L), mean (SD) | 34.0–54.0 | 33.4 (4.8) | 31.6 (5.6) | 33.8 (4.5) | <0.01 |
| Creatine kinase (U/L), median (IQR) | 30.0–310.0 | 100.0 (58.0–172.0) | 118.0 (69.0–223.0) | 99.0 (51.25–154.0) | 0.03 |
| Lactate dehydrogenase (U/L), median (IQR) | 120–150 | 255 (188–359) | 344 (234–469) | 238 (183–333) | <0.01 |
| Glucose (>11.1mmol/L), n (%) | 3.9–6.1 | 44 (15) | 12 (22) | 32 (14) | 0.14 |
| Procalcitonin (ng/mL), median (IQR) | <0.05 | 0.07 (0.05–0.16) | 0.15 (0.09–0.41) | 0.07 (0.04–0.13) | <0.01 |
| C-reactive protein (mg/L), median (IQR) | ≤5.0 | 23.4 (4.1–38.0) | 34.6 (10.6–71.7) | 19.5 (4.0–37.0) | 0.02 |
| Hyperkalemia (>5.0mmol/L), n (%) | ≤5 | 22 (8) | 10 (18) | 12 (5) | <0.01 |
| Other respiratory pathogens,<sup>1</sup> n (%) | NA | 4 (1) | 0 (0) | 4 (2) | >0.99 |
| CT score,<sup>2</sup> median (IQR) | NA | 7 (5–12) | 9 (6–15) | 7 (5–11) | <0.01 |

AKI, acute kidney injury; CT, computed tomography; IQR, interquartile range; SD, standard deviation
Data are n (%), mean (SD) and median (IQR). P values are comparing AKI patients and non-AKI patients. COVID-19 = 2019 novel coronavirus. NA=not applicable.
<sup>2</sup>The PCR test tested for influenza A virus, influenza B virus, rhinovirus/enterovirus, respiratory syncytial virus A/B, adenovirus, *Mycoplasma pneumoniae*, parainfluenza viruses, *Bordetella pertussis*, *Legionella pneumophila*, and *Chlamydia pneumoniae*.
<sup>2</sup>For each patient, the initial CT scans were evaluated and a quantitative score was assigned based on characteristics reported in recently published literature [9].

The nucleic acid assay for 10 respiratory pathogens antibodies, detected 2 patients with respiratory syncytial virus A/B, 1 patient with *Mycoplasma pneumoniae*, and 1 patient with *Chlamydophila pneumoniae* coinfection. None of the 4 patients with respiratory coinfections developed AKI (**Table 2**). A series of typical CT images of a patient from initial onset of symptoms to recovery is shown in **Fig. 1**. According to chest CT, AKI patients had a significantly higher median CT score that non-AKI patients (**Table 2**).

**Figure 1.**
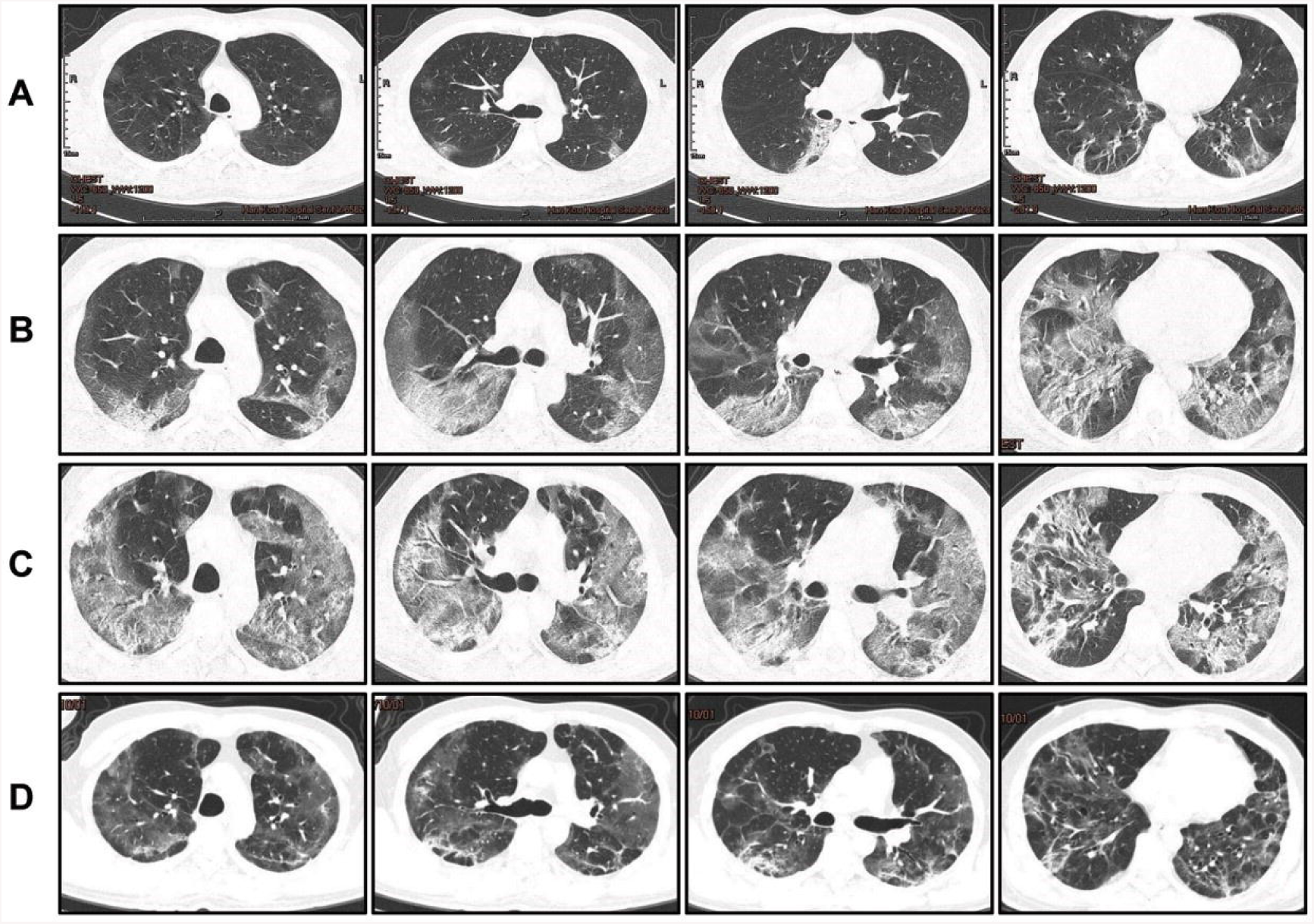
Transverse chest computed tomography (CT) images of the lungsof a 47-year-old man with COVID-19. (A) Bilateral ground glass opacities are visible in the lungs on Day 6 after symptom onset; (B) Diffuse bilateral areas of ground-glass opacities and consolidation of the peripheries of both lungs on Day 11 after symptom onset; (C) Increased diffuse bilateral areas of ground glass opacities and consolidation on Day 18 after symptom onset; (D) Gradual resolution of consolidation and partially reticular pulmonary fibrosis on Day 31 after symptom onset.

Of the 287 patients in the study, 65.5% were discharged by March 8, 2020 (the date on which follow-up was censored). The mortality was 6.6% (**Table 3**). The incidence of stage 1 and stages 2/3 AKI was 14.3% and 4.9%, respectively. The AKI stage was related to the clinical endpoint of the patient. A comparison of outcomes according to AKI status and stage found that, comparing to non-AKI patients and stage 1 and stages 2/3 AKI had significantly lower discharge rates (72.4%, 41.5%, and 14.3% for non-AKI, AKI stage 1, and AKI stages 2/3, respectively) and significantly higher mortality rates (3.0 %, 7.3%, and 64.3% for non-AKI, AKI stage 1, and AKI stages 2/3, respectively). The median length of hospitalization was 18.0, 22.5, and 12.0 days for non-AKI, AKI stage 1, and AKI stages 2/3 patients, respectively.

**Table 3.** Clinical outcomes of COVID-19 patients at data cutoff

|  | All patients<br>(n=287) | Non-AKI<br>(n=232) | AKI stage 1<br>(n=41) | AKI stages 2<br>& 3 (n=14) | *P*<br>Value |
| --- | --- | --- | --- | --- | --- |
| Discharge from hospital, n (%) | 188 (65.5) | 168 (72.4) | 17 (41.5) | 2 (14.3) | <0.01 |
| Hospitalization, n (%) | 80 (27.9) | 57 (24.6) | 21 (51.2) | 3 (21.4) |  |
| Death, n (%) | 19 (6.6) | 7 (3.0) | 3 (7.3) | 9 (64.3) |  |
| Lengths of hospital stay(d), median (IQR) | 18.0<br>(12.0–23.0) | 18.0<br>(13.0–23.0) | 22.5<br>(8.3–28.0) | 12.0<br>(4.5–17.8) | 0.02 |

Data are n (%) and median (IQR).
*P* values are comparing AKI patients and non-AKI patients. *P* <0 .05 was considered statistically significant.

## Discussion

A recent seminal work indicated that the COVID-19 uses the angiotensin converting enzyme II (ACE2) as a cell entry receptor, a cellular mechanism identical to that of the SARS-CoV[14, 15]. In addition, a gain-of-function experiment found that the ACE2 could be used by a wildtype coronavirus of a bat origin to gain entry into cells[15, 16]. ACE2 is not exclusively expressed in the respiratory organs but also in other human tissues. Of note, the expression of ACE2 is almost 100-fold higher in kidney tissue than in the lung, suggesting that kidney cells may be target of infection by SARS-CoV andSARS-CoV-2[15]. An ongoing case study by Li et al. [15], on kidney function in patients with COVID-19 has shown that 63% (32/51) of patients exhibited proteinuria, and 19% (11/59) and 27% (16/59) of the patients had an elevated level of plasma creatinine and blood urea nitrogen, respectively. Moreover, CT scans showed radiographic abnormalities of the kidneys of all the patients[15], and most importantly, SARS-CoV-2 nucleic acids have been found in the urine of patients with COVID-19 [2]. Together, these multiple lines of evidence indicate that kidney impairment may be one of the major causes of morbidity among patients with COVID-19, and may contribute to multiorgan failure and death.

Our study investigated the incidence of AKI and risk factors for AKI in patients with COVID-19. The study included 287 patients admitted from Hankou Hospital, Wuhan from February 8 to March 8, 2020. The incidence of AKI among the patients in our study was substantially higher than the overall incidence of 0.5-7% reported in previous studies[2, 7], and is comparable to that of critically ill patients hospitalized with other illnesses (29%) [3]. The following reasons may explain the higher incidence of AKI observed in our study compared to previous studies: First, most of the patients admitted to our hospital with COVID-19 had severe disease. At the time of admission, 43% of the patients had severe disease. Although many patients had low pulse oxygen saturation when they were admitted to the hospital, the symptoms of dyspnea tended to be mild. The reason might be that the patient’s condition gradually deteriorated and the body adapted and compensated to some degree. The very high CT score and the rapid deterioration of many patients after admission also provide evidence of the severity of the disease among the patients in this study. Second, the methods that we used to diagnose and stage AKI might have overestimated the incidence and severity. Due to the outbreak of COVID-19 and the shortage of medical resources, many patients did not have a baseline serum creatinine test prior to admission, and had not had a creatinine test in the previous year. Therefore, we could only estimate the baseline serum creatinine of such patients according to the patient’s sex, weight, medical history and other related parameters. The incidence of AKI was determined based on baseline values and serum creatinine values within 7 days of admission. In order to reduce the bias of our estimates of baseline values, the two methods, including average serum creatinine method [11] and eGFR 75 method [17], were used for comparison. The AKI incidence rate was 25.8% according to the average serum creatinine method and 19.2% according to the eGFR75 method (**Supplemental Table 2**). The calculated values of baseline serum creatinine estimated by the eGFR75 method were significantly different from those of some of the patients with baseline serum creatinine results available. Imprecision and inaccuracies in estimated baseline value of serum creatinine might have caused an overestimation of the incidence of AKI in our study.

However, any overestimation of the incidence of AKI may not have directly affected our investigation of AKI risk factors. We found that compared with non-AKI patients with COVID-19, older age, multiple pre-existing comorbidities, an increased white blood cell count, low lymphocyte count, and increased levels of PCT and CRP, were risk factors for AKI. The prevalence of several of the pre-existing comorbidities that we considered, including chronic lung diseases and diabetes mellitus, did not differ significantly between the patients with and without AKI, although this might be due to the relatively small number of cases of AKI in this study. Lymphopenia indicates that decreased immunity might occur in patients with COVID-19, and an increased level of PCT and CRP suggest that some patients may have had secondary bacterial infections which caused excessive inflammatory responses. Unfortunately, we did not conduct bacterial and fungal culture on the patients with respiratory coinfections detected on PCR because of the shortage of medical staff, and were unable to test patients’ inflammatory factor profiles because the local hospital did not have inflammatory factor test kits. Previous studies [2–4, 7] have shown that patients with COVID-19 have significantly increased D-dimer levels, multiple organ damage, and electrolyte disturbances. Our study revealed that these abnormalities were more pronounced in patients with AKI. These results suggest that patients may experience severe hypoxia, resulting in aggravated multiple organs damage. It should not be overlooked that SARS-CoV-2 might also directly attack multiple organs, especially the kidneys. The occurrence and development of AKI often leads to the imbalance of blood volume and electrolytes, the accumulation of metabolites, the aggravation of multiple organ dysfunction, and creating a vicious cycle. While, the majority of patients with stage 1 AKI recovered and were discharged, those who progressed to stage 2/3 AKI, had an extremely high mortality rate. The underlying reason might be due to the limited treatment conditions in urgent breakout period of our country. The study results suggest that early intervention after the occurrence of AKI is very important, and that continuous renal replacement should be considered to help alleviate the progress of AKI.

Our survey included all clinically confirmed COVID-19 cases because most suspected cases (also known as clinically confirmed cases) present an exact epidemiological history, and many patients changed from mild to severe during home quarantine. Due to the long observation time, many patients showed negative by RT-PCR assay when they were hospitalized. In fact, there are still a few suspected cases turned positive after with two negative RT-PCR results and discharged, which should be recognized as laboratory-confirmed cases. In addition, other laboratory tests such as infection-related indicators (PCT and CRP), common respiratory pathogen nucleic acid determination, and CT scanning can also help us to differentiate pneumonia.

Our research has the following shortcomings: First, due to the large number of patient populations and the shortage of medical staff and equipment, we did not perform blood gas analysis and blood lactate measurement on most patients. Instead, we based on the patient’s main complaint, pulse oxygen saturation monitoring and CT to indirectly judge the oxygenation. Second, we have not explored the impact of treatment on AKI, because our treatments are based on WHO’s standardized procedure [18]. The antiviral drugs, antibacterial drugs and hormones have little effect on renal function. Third, we did not exclude patients with chronic renal insufficiency. Five patients had chronic renal insufficiency with 3 cases in AKI group and 2 in the non-AKI group. These patients were related to hypertension (2 cases) and diabetes (3 cases) at admission, but showed normal creatinine level during the past year. Fourth, due to the urgent collation of data, we have a shorter follow-up time for some patients, and the shortest hospital stay for patients is only over 14 days. These may interfere with the final prognosis judgment and lead to the failure to analyze the survival time. Fifth, we did not analyze the comorbidities of all patients such as shock and adult respiratory distress syndrome (ARDS), because our focus is to analyze the risk factors of AKI that were present on admission in order to provide evidence for the development of COVID-19 prevention and treatment guidelines.

## Conclusion

Kidney is a primary target organ of SARS-CoV-2 and the incidence of AKI is high in patients hospitalized with COVID-19. Deterioration of kidney function aggravates other organ damage. We identified risk factors for AKI in patients with COVID-19. These included older age, severe pneumonia, and pre-existing cardiovascular and renal disease. Although the incidence of stage 1 AKI in critically ill patients is high, most of them recover. However, patients who progress to stage 2/3 AKI have a very high mortality rate. Thus, the prevention of AKI and monitoring of kidney function is very important in the clinical management of COVID-19 patients. Future studies of AKI in COVID-19 patients should obtain more accurate baseline creatinine values and use urine volume measurements as the basis for AKI determination. In addition, larger, multi-center studies are needed to gain a better understanding of AKI in individuals with COVID-19, and how to prevent and manage it.

## Data Availability

Data sharing is not applicable to this article as no datasets were generated or analysed during the current study.

## Acknowledgments

This work was supported by the Natural Science Foundation of China Grant 81871604; the Natural Science Foundation of Guangdong Province, China, Grants 2016A030310389 and 2017A030313590; the Outstanding Youths Development Scheme of Nanfang Hospital, Southern Medical University, Guangzhou, China, Grant 2016J011.

## Declarations

### Ethics approval and consent to participate

This study was approved by the National Health Commission of China and the Ethics Commission of Hankou Hospital (hkyy2020-005). The Ethics Commission of Hankou Hospital waived the requirement for obtaining informed consent.

### Consent for publication

Not applicable.

### Authors’ contributions

Z.Z. and Z.C. contributed substantially to the conception and design of the study, Q.H. and Z.Z. drafted and provided critical revision of the article. G.X. and F.W. acquired the primary data, H.H. and T.S. analyzed and interpretation of the data. H.L., J.H., and W.S. analyzed the imaging data. All the authors provided final approval of the version submitted for publication.

## List of abbreviations

AKI: Acute kidney injury
COVID-19: Coronavirus disease 2019
eGFR: Estimated glomerular filtration rate
MDRD: Modification of Diet in Renal Disease
CT: Computed tomography
PCR: Polymerase chain reaction
ACE2: Angiotensin converting enzyme II
SARS-CoV: Serve acute respiratory syndrome coronavirus
WHO: World Healthy Organization

## Additional files

*Supplemental Table 1.doc*: Other symptoms of COVID-19 patients on admission *Supplemental Table 2*.*doc*: The accuracy of AKI diagnosis between the average SCr method and the eGFR 75 method

